# Influence of nursing staff working hours on the stress level during the COVID-19 pandemic: a cross-sectional online survey

**DOI:** 10.1101/2020.08.12.20173385

**Authors:** Manuela Hoedl, Silvia Bauer, Doris Eglseer

## Abstract

**Background:** Working as a nurse means being responsive and highly accountable 24/7 and to be able to offer high-quality care, specifically during pandemics. Studies have shown that the average number of working hours per week is a significant predictor of stress and that the severity of the COVID-19 pandemic has increased the nurses’ stress levels.

**Objective:** Therefore, we investigated (1) if a change had occurred in the nurses’ working hours during the COVID-19 pandemic as compared to the hours employed and (2) the influence of the nursing staff’s working hours during COVID-19 pandemic on the perceived level of stress.

**Design:** We used an online survey in this Austrian cross-sectional study, distributed using a snowball sampling method.

**Participants and methods:** In the online survey, we asked the nurses many relevant questions, including how many hours they are employed per week and how many hours they had worked on average per week since the outbreak of COVID-19. We used the Perceived Stress scale to measure stress level among these nurses. Data were collected between mid-May and mid-July 2020.

**Results:** Three-quarters of the 2600 participating nurses reported changes in their working hours during the COVID-19 pandemic. The nursing staff’s hours of employment were statistically significantly associated with their average number of working hours during the COVID-19 pandemic. About two-thirds of the nurses who were employed either less than 10 hours or for 31-40 hours worked for more than 40 hours. Most of the nurses experienced a moderate level of stress. We identified a statistically significant association between increasing the number of working hours per week and the nurses’ perceived stress level. In addition, 15% of the nurses who had worked more than 40 hours reported experiencing a high level of stress. In addition, we found that nurses who worked more hours during the pandemic experienced higher stress than nurses who reduced their working hours or kept the same working hours.

**Conclusions:** We found a statistically significant association between an increase in the nursing staff’s working hours and their level of stress. We believe that these results reflect the negative consequences of prolonged working hours. For this reason, a (inter-)national discussion is needed on the topic of restricting the working hours of healthcare workers during such pandemics. This discussion can improve the health and safety of the health care workers, the patients, as well as members of the general population.

**Tweetable Abstract:** Increasing working hours of nurses during COVID-19 pandemic is increasing their level of stress.

What is already known about the topic?
- The ability of the nurse to provide high-quality care is strongly associated with the health of nursing staff
- During COVID-19 pandemic, prolonged/irregular working hours may be appearing, to ensure continuity of the healthcare system.
- Such prolonged/irregular working hours can result in increased stress among nurses.

What this paper adds
- Three-quarters of the nurses reported changes in their working hours during the COVID-19 pandemic.
- About two-thirds of the nurses who were employed either less than 10 hours or for 31–40 hours worked for more than 40 hours.
- This study showed that an increasing number of working hours per week is increasing nurses’ perceived stress level.
- It also demonstrated that 15% of the nurses who had worked more than 40 hours experienced a high level of stress.

## Background

The healthcare system has been organised to ensure that 24-hour-a-day services are provided seven days a week (24/7). Within the healthcare system, nurses represent the largest professional group; therefore, the overall quality of the healthcare system largely depends on the nurses’ performance. Nevertheless, the ability of the nurse to provide high-quality care is strongly associated with the health of nursing staff (Rosa et al., 2018).

To work as a nurse – either as a qualified nurse or nursing aide – individuals need to be responsive and highly accountable 24/7 in order to fullfill the health service requirements (Rosa et al., 2018). Therefore, nurses need to have worked for an adequate amount of time in the health sector to be able to provide high-quality care (Rosa et al., 2018). In the European Union, the 2003 Working Time Directive sets limits of 48 hours per week, an amount of working time that includes overtime, calculated as an average over a maximum of four months (European Parliament & Union, 2003). This directive also defines a daily amount of rest, i.e. a minimum 11 hours, and a weekly amount of rest, i.e. normally 24 hours plus the 11-hour daily rest period and the amount of time spent performing work at night (European Parliament & Union, 2003).

However, the directive allows deviations/exemptions in Article 17 paragraph 3 (European Parliament & Union, 2003); these deviation/exemptions were required during the recent COVID-19 pandemic. This might have led to prolonged/irregular working hours for nursing staff. Such prolonged/irregular working hours can include working periods that exceed the conventional working hours, overtime, shift work, night work and on-call scheduling.

These long/irregular hours worked by nurses can have negative consequences on the health and safety of both nurses and their patients. Several studies have shown that such prolonged/irregular working hours can results in nurses having a lower ability to detect adverse changes in their patients or address them in a timely manner (A. M. Trinkoff et al., 2011), reduced or lacking patient safety, fair to poor quality of care offered, more care activities left undone (Peter Griffiths et al., 2014), or adverse nursing outcomes such as complaints from patients or families (Son et al., 2019).

Another study reported several negative consequences for the nurses, such as increased emotional and mental fatigue, the disruption of normal sleeping and waking hours, depression and various illnesses such as musculoskeletal disorders (Harris et al., 2015). These disruptions could also result in a diminished capacity to manage the workload, job dissatisfaction, burn-out, absenteeism and poor service delivery and productivity (Aiken et al., 2002; Conway et al., 2008; Rajbhandary & Basu, 2010). Another recent systematic review reported that working 55 hours or more per week increases the risk of stroke (Descatha et al., 2020).

Furthermore, irregular work schedules and work overload can lead to professional and personal conflicts, which may affect personal and family responsibilities in the long run and decrease the nurses’ levels of satisfaction with their work and lives (Messenger & Vidal, 2015; Yildirim & Aycan, 2008). Finally, these prolonged/irregular working hours also affect the absenteeism rates, recruitment and retention of nursing staff (Messenger & Vidal, 2015; Yildirim & Aycan, 2008).

Two more recent studies have been carried out to investigate the level of stress of nurses. Whereas Almazan et al. concluded that the average number of working hours per week was a significant predictor of stress (Almazan et al., 2019). Another study reported that high stress can lead to a decline in the quality of nursing care (Keykaleh et al., 2018). In addition, caring for COVID-19 affected persons or suspected cases might also be influencing the level of stress experienced by nurses during this pandemic. As an example, authors of a recent study showed that the perceived severity of the COVID-19 pandemic increases the stress level of the nurses (Mo et al., 2020).

Overall, the consequences of nurses working prolonged/irregular working hours are enormous, and also result in increased stress among nurses. Caring for COVID-19-affected persons or suspected cases might also influence the nurses’ level of stress. Even though the European Union has a working time directive, this can be legally overridden during the COVID-19 pandemic, in order to ensure the continuity of the healthcare service. Therefore, we investigated (1) if a change had occurred in the nurses’ working hours during the COVID-19 pandemic as compared to the hours listed in their employment contract and (2) the influence of the nursing staff’s working hours during COVID-19 pandemic on the perceived level of stress.

## Methods

### Design

For this cross-sectional study, we used an online questionnaire, which was distributed using a snowball sampling method and with the aid of social media. The online survey was opened on 12 May 2020 and closed on 13 July 2020.

### Setting and Sample

The authors invited Austrian nursing staff, including nurses, nursing aids and care aids who were working at the bedside with patients/residents during the COVID-19 pandemic. We invited nursing staff working in different settings, such as hospital and long-term care facilities. Nursing leaders, managers, or directors were not invited to take part in the survey, so that we could obtain detailled insight into nursing practice during this pandemic.

### Data collection instrument

We collected demographic sample characteristics such as the year of birth and gender. In addition, we asked the nurses how many hours they were employed per week and to estimate the average number of hours they had worked per week since the outbreak of the COVID-19 pandemic in Austria (i.e. mid-March 2020). Possible answers to this question were: < 10 hours, 10-20 hours, 21-30 hours, 31-40 hours, or > 40 hours.

On a structural level, we collected data on the perceived change in the working hours due to the COVID-19 pandemic (possible answers: Yes, working hours were prolonged; Yes, working hours were reduced; Yes, working hours were changed; No change).

To assess the perceived stress level among the nurses, we used the German version of the Perceived Stress Scale (PSS) on the outcome level (Cohen et al., 1994; Schneider et al., 2017). The PSS has good psychometric properties (Cohen et al., 1994) and consists of ten items, which highlighs its practicability (Klein et al., 2016). The PSS scores can range from 0 to 40, with higher scores indicating higher perceived stress. In addition, categories can be constructed with scores ranging from 0-13 to indicate a low perceived stress level; 14-26, moderate perceived stress level; and 27-40, high perceived level of stress (Cohen et al., 1994; Schneider et al., 2017)..

The PSS is a frequently used instrument for assessing stress in nursing staff (Aalaa et al., 2017; Cicolini et al., 2016; dos Santos et al., 2016; Montanari et al., 2019; Peters et al., 2020; Rayan et al., 2019; Uzun & Sevinc, 2015; Wright, 2018).

### Data analysis

We excluded three participants from this analysis, because they were 69-70 years of age, which is older than the retirement age in Austria.

We calculated descriptive statistics, including percentages for categorical variables and means and a standard deviation for metric variables. We used the X^2^ test to assess categorical variables and, depending on the number of answer categories, we used Cramer (V) or the Contingency Coefficient (CC) as a measure of the effect size. In order to validate our findings with regard to the influence of changes in the working hours on the PSS sum, we conducted an independent-samples Kruskal-Wallis test. We set the p-values < .05 as statistically significant.

### Ethics

The ethical committee of the Medical University of Graz approved the study (32-386 ex 19/20). All participants had to click on „Yes” if they wanted to participate in the survey. The data collection was anonymous, and the collected data were stored on the Medical University of Graz server.

## Results

Table 1 displays the sample characteristics with regard to demographics, number of working hours employed, if a change in working hours during the COVID-19 occurred and if nurses were caring for a suspected case or a person infected with COVID-19.

**Table 1.** Sample characteristics

|  | Nursing staff (*N* = 2602) |
| --- | --- |
| Female % | 83.7 |
| Mean age in years (SD) | 38 (11) |
| Number of hours per week employed % |  |
| < 10 hours | 3.3 |
| 10-20 hours | 8.8 |
| 21-30 hours | 16.6 |
| 31-40 hours | 71.4 |
| Change in working hours % |  |
| Yes, prolonged | 13.3 |
| Yes, reduced | 4.1 |
| Yes, changed, e.g. through reorganisation | 58.3 |
| No changes | 24.4 |
| Caring for a suspected case or a person infected with COVID-19 % |  |
| Yes | 66.5 |
| No | 33.5 |

2602 nurses participated in the survey, most of whom were employed between 31 and 40 hours per week (Table 1). Of this number, 75.6% of the nurses reported changes in their working hours during the COVID-19 pandemic. Nearly 60% of the nurses described changes, e.g. due to reorganisation, and 13.3% reported that their working hours were prolonged during the COVID-19 pandemic. Two-thirds of these nurses were caring for a suspected case or a person infected with COVID-19 during their working hours.

### Comparison of hours of employment and current working hours during the COVID-19 pandemic

In order to give a detailled insight, Table two displays the hours of employment per week as compared to the average number of working hours per week during the COVID-19 pandemic.

**Table 2:** Comparison of hours of employment per week and average number of working hours per week during the COVID-19 pandemic

| Hours of employment per week* | Average working hours per week during COVID-19 pandemic % |  |  |  |  |
| --- | --- | --- | --- | --- | --- |
|  | < 10 | 10-20 | 21-30 | 31-40 | > 40 |
| < 10 (*n* = 85) | 30.6 | 3.5 | 8.2 | 20.0 | 37.6 |
| 10-20 (*n* = 228) | 5.3 | 64.0 | 19.3 | 5.3 | 6.1 |
| 21-30 (*n* = 431) | 1.6 | 13.5 | 59.2 | 20.2 | 5.6 |
| 31-40 (*n* = 1858) | 2.3 | 2.3 | 8.8 | 53.7 | 33.0 |
| Total (*N* = 2602) | 3.3 | 9.6 | 18.1 | 42.8 | 26.2 |

\* $p < .05$

We identified a statistically significant association between the hours of employment and the average number of working hours during the COVID-19 pandemic among nursing staff (*V* = .487, *p* = .000). About two-thirds of the nurses who were employed less than 10 hours worked more than their hours of employment. Most of the nurses who were employed for more than 10 hours worked the hours for which they were employed. One-third of the nurses that were employed 31-40 hours worked more than 40 hours per week during the COVID-19 pandemic.

### Influence of working hours on stress

**Table 3:** Association between the average number of working hours per week during the COVID-19 pandemic and perceived stress among nursing staff

| Average working hours per week during COVID-19* | PSS categories % |  |  |
| --- | --- | --- | --- |
|  | Low | Medium | High |
| < 10 (*n* = 87) | 35.6 | 58.6 | 5.7 |
| 10-20 (*n* = 249) | 39.4 | 52.6 | 8.0 |
| 21-30 (*n* = 470) | 34.7 | 57.2 | 8.1 |
| 31-40 (*n* = 1113) | 32.5 | 58.1 | 9.3 |
| > 40 (*n* = 683) | 29.0 | 55.6 | 15.4 |
| Total (*N* = 2602) | 32.7 | 56.8 | 10.5 |

\* $p < .05$

The majority of nurses experienced a moderate level of stress. We identified a statistically significant association between an increase in the number of working hours per week and the perceived stress level of the nurses (*V* = .078, *p* = .000). About 15% of the nurses who worked more than 40 hours per week reported experiencing a high level of stress.

In order to give detailled insight into how the change in working hours influenced stress, Table four shows the influence of the change in working hours during the COVID-19 pandemic on the nurses’ stress levels. We defined the working hours as changed when the hours of employment per week and average working hours per week during COVID-19 pandemic differed. Some nurses might have worked less, thesame, or more during the COVID-19 pandemic than their official employment contract stipulated.

**Table 4:** Association between changed working hours during the COVID-19 pandemic and perceived level of stress among nursing staff

| Change in working hours* | PSS categories % |  |  |
| --- | --- | --- | --- |
|  | Low | Moderate | High |
| Reduced ($n = 325$) | 37.5 | 53.5 | 8.9 |
| Same ($n = 1424$) | 34.6 | 56.7 | 8.8 |
| Increased ($n = 853$) | 27.9 | 58.3 | 13.8 |

\* $p < .05$

No matter how the working hours had changed, more than half of the nurses experienced moderate stress (53.5%-58.3%). We found a statistically significant association between the three groups with regard to the perceived stress level (CC = 0.097, *p* = 0.000), meaning that nurses with increased working hours experienced higher stress than nurses with reduced or the same working hours.

In order to validate these findings, we calculated an independent-sample-Kruskal-Wallis test to assess the effect of the change in the working hours on the PSS sumscore. We found a statistically significant increase in nursing staff stress level when the hours increased as compared to when the hours were reduced (p = .001) or stayed the same (p = .000). We found no statistically significant difference in the stress level when we compared nurses who worked reduced or the same working hours (p = .850).

## Disussion

This study was carried out to investigate the influence of nursing staff working hours during the COVID-19 pandemic on the perceived level of stress. More than 2600 nurses participated in the survey, most of whom were employed between 31 and 40 hours per week. Three-quarters of the nurses reported changes in their working hours during COVID-19. We identified a statistically significant association between the hours of employment and the average working hours during the COVID-19 pandemic among nursing staff. About two-thirds of the nurses who were employed less than 10 hours worked more than their hours of employment, and one-third of the nurses that were employed 31-40 hours worked more than 40 hours during the COVID-19 pandemic. The majority of nurses experienced a moderate level of stress. We identified a statistically significant association between an increase in working hours per week and the nurses’ perceived stress levels. In addition, 15% of the nurses who worked more than 40 hours reported experiencing a high level of stress. We also found that nurses with increased working hours experienced higher stress than nurses with reduced or the same working hours.

Our results show that most nursing staff work between 31-40 hours per week. This could be explained by the high number of younger nurses who participated in the study, and who may not yet have family responsibilties. This finding is underpinned by a study which highlighted that 39% of the nursing staff in Austria are younger than 40 years of age (Federal Ministry of Labor, 2019).

We also compared the hours of employment and the average number of working hours during the COVID-19 pandemic. When the actual working hours were compared to the hours of employment, we saw that 25.7% and 37.3% of the nurses worked for more hours than they were legally obliged to.. This is surprising, as the Austrian government recommended in mid-March that all unnecessary health treatments, e.g. knee replacement surgery, should be posponed, which should have decreased the number of hospital patients (Federal Ministry for Social Affairs, 2020).

We identified a statistically significant association between the number of working hours and the level of stress among nursing staff. The majority of nurses who were working up to 40 hours per week experienced a low to moderate level of stress, whereas about 70% of the nurses who worked more than 40 hours reported a high perceived level of stress.

These findings are in line with those of previous studies that linked long/irregular working hours to emotional and mental fatigue, disruptions of normal sleeping and waking hours, depression and various illnesses such as musculoskeletal disorders (Harris et al., 2015). In addition, the health and safety of the patients decreased as the nurses’ working hours (P Griffiths et al., 2014; Son et al., 2019; A. Trinkoff et al., 2011).

This is an especially interesting result in times of a pandemic. If patient-safety indicators such as accidents, near misses, failures, or errors in healthcare occur during a pandemic, this might result in more severe consequences than in “normal” times. In addition, such failures to ensure patient safety could actually increase the effect of the pandemic on the general population, through healthcare workers who (unintentionally) act as coronavirus superspreaders. In any case, these results indicate that a discussion about working hour regulations for healthcare workers during such a pandemic should be initiated, as these might increase the health and safety of the healthcare workers, the patients and members of the general population.

### Limitations

We assessed the nurses’ working hours during the COVID-19 pandemic, which might not reflect the nurses’ workload during normal working hours. Studies have shown that the more patients a nurse has to care for, the higher the intensity of work, which can result in an increased risk of accidents and work-related stress, high levels of sick leave and absenteeism (Messenger & Vidal, 2015) and, consequently, injuries and diseases including fatigue and burnout. Inadequate staffing levels also have implications for patient safety and quality of care in terms of higher morbidity and mortality (Poghosyan et al., 2010; Zhu et al., 2012; Aiken et al., 2014). On the other hand, a recent study concluded that the average number of working hours per week was a significant predictor of stress (Almazan et al., 2019). Another limitation of this study, is that we did not assess other factors, such as the working climate, which might also have an influence on the perceived level of stress and, specifically, in this kind of pandemic. One more limitationis that we used a convenience sample. Therefore, our results might cannot be generalised to they entire country of Austria. However, this initial study conducted during a pandemic provides us with important insights on this topic that encourage us and other researchers to repeat the study with larger sample sizes.

## Conclusions

This study was carried out to investigate the influence of nursing staff working hours during the COVID-19 pandemic on the perceived level of stress. We found a statistically significant association between an increase in working hours and the level of perceived stress among nursing staff.

We believe that these results reflect the negative consequences of prolonged working hours. For this reason, a (inter-)national discussion is needed on the topic of restricting the working hours of healthcare workers during such pandemics. This discussion can improve the health and safety of the health care workers, the patients, as well as members of the general population and can even save lifes.

## Data Availability

Due to legal issues the data can not be made available.

## Author contributions

All above listed as authors (M. Hoedl, S. Bauer & D. Eglseer) are qualified for authorship by meeting all four of the following criteria:

1. Have made substantial contributions to conception and design, or acquisition of data, or analysis and interpretation of data;
2. Been involved in drafting the manuscript or revising it critically for important intellectual content;
3. Given final approval of the version to be published. Each author should have participated sufficiently in the work to take public responsibility for appropriate portions of the content; and
4. Agreed to be accountable for all aspects of the work in ensuring that questions related to the accuracy or integrity of any part of the work are appropriately investigated and resolved.

## Acknowledgments

We would like to thank all participating nurses.

## References

Aalaa, M., Najmi Varzaneh, F., Maghbooli, Z., Samandari, N., Mostafavi, A., Salemi, S., Mehrdad, N., & Sanjari, M. (2017). Influence of MTHFR gene variations on perceived stress modification: Preliminary results of NURSE study. Med J Islam Repub Iran, 31, 128. https://doi.org/10.14196/mjiri.31.128

Almazan, J. U., Albougami, A. S., & Alamri, M. S. (2019, 2019/08/01/). Exploring nurses’ work-related stress in an acute care hospital in KSA. Journal of Taibah University Medical Sciences, 14(4), 376–382. https://doi.org/10.1016/j.jtumed.2019.04.006

Cicolini, G., Della Pelle, C., Cerratti, F., Franza, M., & Flacco, M. E. (2016, Jul). Validation of the Italian version of the Stanford Presenteeism Scale in nurses. J Nurs Manag, 24(5), 598–604. https://doi.org/10.1111/jonm.12362

Cohen, S., Kamarck, T., & Mermelstein, R. (1994). Perceived stress scale. Measuring stress: A guide for health and social scientists, 10.

Descatha, A., Sembajwe, G., Pega, F., Ujita, Y., Baer, M., Boccuni, F., Di Tecco, C., Duret, C., Evanoff, B. A., Gagliardi, D., Godderis, L., Kang, S.-K., Kim, B. J., Li, J., Magnusson Hanson, L. L., Marinaccio, A., Ozguler, A., Pachito, D., Pell, J., Pico, F., Ronchetti, M., Roquelaure, Y., Rugulies, R., Schouteden, M., Siegrist, J., Tsutsumi, A., & Iavicoli, S. (2020, 2020/09/01/). The effect of exposure to long working hours on stroke: A systematic review and meta-analysis from the WHO/ILO Joint Estimates of the Work-related Burden of Disease and Injury. Environment International, 142, 105746. https://doi.org/10.1016/j.envint.2020.105746

dos Santos, T. M., Kozasa, E. H., Carmagnani, I. S., Tanaka, L. H., Lacerda, S. S., & Nogueira-Martins, L. A. (2016, Mar-Apr). Positive Effects of a Stress Reduction Program Based on Mindfulness Meditation in Brazilian Nursing Professionals: Qualitative and Quantitative Evaluation. Explore (NY), 12(2), 90–99. https://doi.org/10.1016/j.explore.2015.12.005

DIRECTIVE 2003/88/EC OF THE EUROPEAN PARLIAMENT AND OF THE COUNCIL of 4 November 2003 concerning certain aspects of the organisation of working time, (2003).

Federal Ministry for Social Affairs, H., Care and Consumer Protection,. (2020, 12. March 2020). Empfehlung zur Verschiebung aller nicht vordringlichen Untersuchungen und Behandlungen

Federal Ministry of Labor, S. A., Health and Consumer Protection;. (2019). Pflegepersonal-Bedarfsprognose für Österreich.

Griffiths, P., Dall’Ora, C., Simon, M., Ball, J., Lindqvist, R., Rafferty, A.-M., Schoonhoven, L., Tishelman, C., Aiken, L. H., & Consortium, R. C. (2014). Nurses’ shift length and overtime working in 12 European countries: the association with perceived quality of care and patient safety. Medical Care, 52(11), 975–981. https://doi.org/10.1097/MLR.0000000000000233

Griffiths, P., Dall’Ora, C., Simon, M., Ball, J., Lindqvist, R., Rafferty, A. M., Schoonhoven, L., Tishelman, C., Aiken, L. H., & & Consortium, R. C. (2014). Nurses’ shift length and overtime working in 12 European countries: the association with perceived quality of care and patient safety. Medical Care, 52(11), 975–981. https://doi.org/10.1097/MLR.0000000000000233

Harris, R., Sims, S., Parr, J., & Davies, N. (2015, Feb). Impact of 12h shift patterns in nursing: a scoping review. Int J Nurs Stud, 52(2), 605–634. https://doi.org/10.1016/Minurstu.2014.10.014

Keykaleh, M. S., Safarpour, H., Yousefian, S., Faghisolouk, F., Mohammadi, E., & Ghomian, Z. (2018). The relationship between nurse’s job stress and patient safety. Open access Macedonian journal of medical sciences, 6(11), 2228. https://doi.org/10.3889/oamjms.2018.351

Klein, E. M., Brahler, E., Dreier, M., Reinecke, L., Muller, K. W., Schmutzer, G., Wolfling, K., & Beutel, M. E. (2016, May 23). The German version of the Perceived Stress Scale - psychometric characteristics in a representative German community sample. BMC Psychiatry, 16, 159. https://doi.org/10.1186/s12888-016-0875-9

Messenger, J. C., & Vidal, P. (2015). The organization of working time and its effects in the health services sector: a comparative analysis of Brazil, South Africa and the Republic of Korea. https://EconPapers.repec.org/RePEc:ilo:ilowps:994869453402676

Mo, Y., Deng, L., Zhang, L., Lang, Q., Liao, C., Wang, N., Qin, M., & Huang, H. (2020, Apr 7). Work stress among Chinese nurses to support Wuhan for fighting against the COVID-19 epidemic. J Nurs Manag. https://doi.org/10.1111/jonm.13014

Montanari, K. M., Bowe, C. L., Chesak, S. S., & Cutshall, S. M. (2019, Jun). Mindfulness: Assessing the Feasibility of a Pilot Intervention to Reduce Stress and Burnout. J Holist Nurs, 37(2), 175–188. https://doi.org/10.1177/0898010118793465

Peters, A., El-Ghaziri, M., Quinn, B., Simons, S., & Taylor, R. (2020, Jan 8). An Exploratory Study of Bullying Exposure Among School Nurses: Prevalence and Impact. J Sch Nurs, 1059840519897308. https://doi.org/10.1177/1059840519897308

Rayan, A., Sisan, M., & Baker, O. (2019, Jun). Stress, Workplace Violence, and Burnout in Nurses Working in King Abdullah Medical City During Al-Hajj Season. Journal of Nursing Research, 27(3), e26. https://doi.org/10.1097/jnr.0000000000000291

Rosa, J. D., Postic, N., Wiskow, C., & Humblet, M. (2018). Health Services - Decent Working Time for Nursing Personnel: Critical for Worker Well-being and Quality Care (Policy brief: Health Services, Issue. I. L. Office. https://www.ilo.org/sector/Resources/publications/WCMS_655277/lang--en/index.htm

Schneider, E. E., Schönfelder, S., Wolf, M., & Wessa, M. (2017, 09/01). All stressed out? Introducing a German version of the perceived stress scale: Validation, psychometric properties and sample differences in healthy and clinical populations. Psychoneuroendocrinology, 83, 21. https://doi.org/10.1016/j.psyneuen.2017.07.296

Son, Y.-J., Lee, E. K., & Ko, Y. (2019). Association of Working Hours and Patient Safety Competencies with Adverse Nurse Outcomes: A Cross-Sectional Study. Int J Environ Res Public Health, 16(21), 4083. https://doi.org/10.3390/ijerph16214083

Trinkoff, A., Johantgen, M., Storr, C., Gurses, A., Liang, Y., & Han, K. (2011, 01/01). Nurses’ Work Schedule Characteristics, Nurse Staffing, and Patient Mortality. Nurs Res, 60, 1–8. https://doi.org/10.1097/NNR.0b013e3181fff15d

Trinkoff, A. M., Johantgen, M., Storr, C. L., Gurses, A. P., Liang, Y., & Han, K. (2011, Jan-Feb). Nurses’ work schedule characteristics, nurse staffing, and patient mortality. Nurs Res, 60(1), 1–8. https://doi.org/10.1097/NNR.0b013e3181fff15d

Uzun, O., & Sevinc, S. (2015, Dec). The relationship between cultural sensitivity and perceived stress among nurses working with foreign patients. J Clin Nurs, 24(23-24), 3400-3408. https://doi.org/10.1111/jocn.12982

Wright, E. M. (2018, Jun). Evaluation of a Web-Based Holistic Stress Reduction Pilot Program Among Nurse-Midwives. J Holist Nurs, 36(2), 159–169. https://doi.org/10.1177/0898010117704325

Yildirim, D., & Aycan, Z. (2008, 03/01). Nurses’ work demands and work-family conflict: A questionnaire survey. Int J Nurs Stud, 45, 1366–1378. https://doi.org/10.1016/j.ijnurstu.2007.10.010

